# How best do we engage the general population in testing for COVID-19?

**DOI:** 10.1101/2021.01.29.21250730

**Authors:** Daniella Watson, Natalia Laverty Baralle, Jawahr Alagil, Krithika Anil, Sandy Ciccognani, Rachel Dewar-Haggart, Sarah Fearn, Julia Groot, Kathryn Knowles, Claire Meagher, Carmel McGrath, Sarah Muir, Jo Musgrove, Kate Glyn-Owen, Kath Woods-Townsend, Andrew Mortimore, Paul Roderick, Janis Baird, Hazel Inskip, Keith Godfrey, Mary Barker, on behalf of Southampton COVID-19 Testing Pilot Programme

**Affiliations:** Global Health Research Institute, School of Human Development and Health, Faculty of Medicine, University of Southampton, United Kingdom; School of Cancer Sciences, Faculty of Medicine, University of Southampton, United Kingdom; School of Health Sciences, Faculty of Life and Environmental Sciences, University of Southampton, United Kingdom; College of Applied Medical Sciences, Health Rehabilitation Department, King Saud University, Kingdom of Saudi Arabia; School of Health Professions, Faculty of Health and Human Sciences, University of Plymouth, Plymouth, UK; School of Primary Care, Population Sciences and Medical Education, Faculty of Medicine, University of Southampton, United Kingdom; School of Human Development and Health, Faculty of Medicine, University of Southampton, United Kingdom; Department of Psychology, University of Bath, United Kingdom; NIHR Southampton Biomedical Research Centre, University of Southampton and University Hospitals Southampton NHS Foundation Trust, United Kingdom; Southampton Education School, Faculty of Social Sciences, University of Southampton, Southampton, United Kingdom; MRC Lifecourse Epidemiology Unit, University of Southampton, United Kingdom; School of Public Health, Faculty of Health Sciences, University of the Witwatersrand, Johannesburg, South Africa

## Abstract

The UK Scientific Advisory Group for Emergencies (SAGE) emphasises the need for high levels of engagement with communities and individuals to ensure the effectiveness of any COVID-19 testing programme. A novel pilot health surveillance programme to assess the feasibility of weekly mass RT-LAMP testing for the SARS-CoV-2 virus using saliva samples collected at home was developed and piloted by the University of Southampton and Southampton City Council. Rapid qualitative evaluation was conducted to explore experiences of those who took part in the programme, of those who declined and of those in the educational and healthcare organisations involved in the pilot testing who were responsible for roll-out. This included 77 interviews and 20 focus groups with 223 staff, students, pupils and household members from four schools, one university, and one community healthcare NHS trust. Conversations revealed that high levels of communication, trust and convenience were necessary to ensure people’s engagement with the programme. This suggests community leaders and stakeholder organisations should be involved throughout programme development and implementation to optimise these features of the testing. Participants’ and stakeholders’ motivations, challenges and concerns need to be understood and these insights used to modify the programme in a continuous, real-time process to ensure and sustain engagement with testing over the extended period necessary.

## Introduction

Universal, repeated, weekly testing followed by strict household isolation after a positive test and continued normal life after a negative test, has been promoted in the UK as an important exit strategy from the current COVID-19 (SARS-CoV-2) pandemic.^(1)^ A novel pilot programme to assess the feasibility of weekly mass RT-LAMP testing^(2,3)^ for the SARS-CoV-2 virus using saliva samples collected at home was developed and piloted in a unique partnership (the Southampton COVID-19 Saliva Testing Programme) between Southampton City Council and the University of Southampton. Between June and October 2020, participants from two general practices in Southampton, staff and students at the University of Southampton and staff and pupils from one infant, one junior, one primary and one secondary school in the city were invited to participate in two phases of a pilot of the Southampton COVID-19 Saliva Testing Programme. The four schools had catchment areas in more deprived parts of the city, and the pupils were from diverse ethnic backgrounds. More than a quarter of students at the University of Southampton come from outside the UK. Interactive engagement activities took place alongside the testing programme to maximise uptake of saliva testing, with extensive development and deployment of educational materials targeting different age groups of students and pupils in the schools, alongside work to engage university students.

The UK Scientific Advisory Group for Emergencies (SAGE) emphasises the need for high levels of engagement with communities and individuals in the creation of successful mass testing programmes.^(4)^ High levels of engagement with the community builds trust, shared goals, and a sense of fairness;^(5, 6)^ engagement also bridges cultural and language gaps.^(7)^ A rapid review of the role of community engagement in the response to COVID-19 found that local community leaders and stakeholders took on important programme functions, including designing and planning, building trust, negotiating community entry and behaviour change and risk communication.^(8)^Rapid qualitative evaluation can be used when the public sector requires immediate feedback on programme process and impacts in order to shape policy.^(9)^ Hence, this type of approach lends itself to providing insights into the response to public health recommendations and programmes during the COVID-19 pandemic. Data generated describing people’s lived experiences of the infection, their behaviour and responses to the government efforts to contain the virus have complemented and explained the epidemiological data.^(10)^ Other studies involving rapid qualitative methods during the pandemic have provided ‘real-time’ feedback on current public health practices^(11)^ and have informed the design of the UK’s COVID-19 response.^(12)^ This approach was established to inform iterative development and testing of programmes and interventions and has been shown to produce findings comparable to those of more traditional qualitative research and thematic analysis.^(13, 14)^

To generate insights and inform the design and modification of the Southampton COVID-19 Saliva Testing Programme, we conducted a rapid qualitative evaluation exploring the experiences of individuals and organisations who took part and of those individuals who declined to take part to inform the development of the next phase of the programme for mass-testing.

### The rapid qualitative evaluation

Between 4^th^ June and 7^th^ November 2020, we conducted 77 interviews and 20 focus groups with 223 staff, pupils, students and household members, from four schools, one university, and two GP practices belonging to one Clinical Commissioning Group in the city of Southampton in the South-East of England. Three groups of participants took part in interviews and focus groups: i) individuals who had been approached about taking part in the pilot Saliva Testing Programme including some who tested positive; ii) those who were approached but declined to take part; and iii) senior university, primary care and school representatives responsible for delivering the Saliva Testing Programme in their organisations. The majority of participants in this qualitative study were recruited via the testing programme’s online registration process, a feedback form on the Southampton City Council website, during a phone call with the testing programme team to those who tested positive or inconclusive, or through the schools’ weekly newsletters about the programme. Senior university and school representatives and a minority of other participants were recruited through the testing team’s and participants’ professional networks. Participants gave permission for their contact details to be shared with the qualitative evaluation team and were offered a small financial incentive to take part. Participants aged under 18 years and their parents signed consent forms. Participants’ characteristics are outlined in Table 1.

**Table 1.**
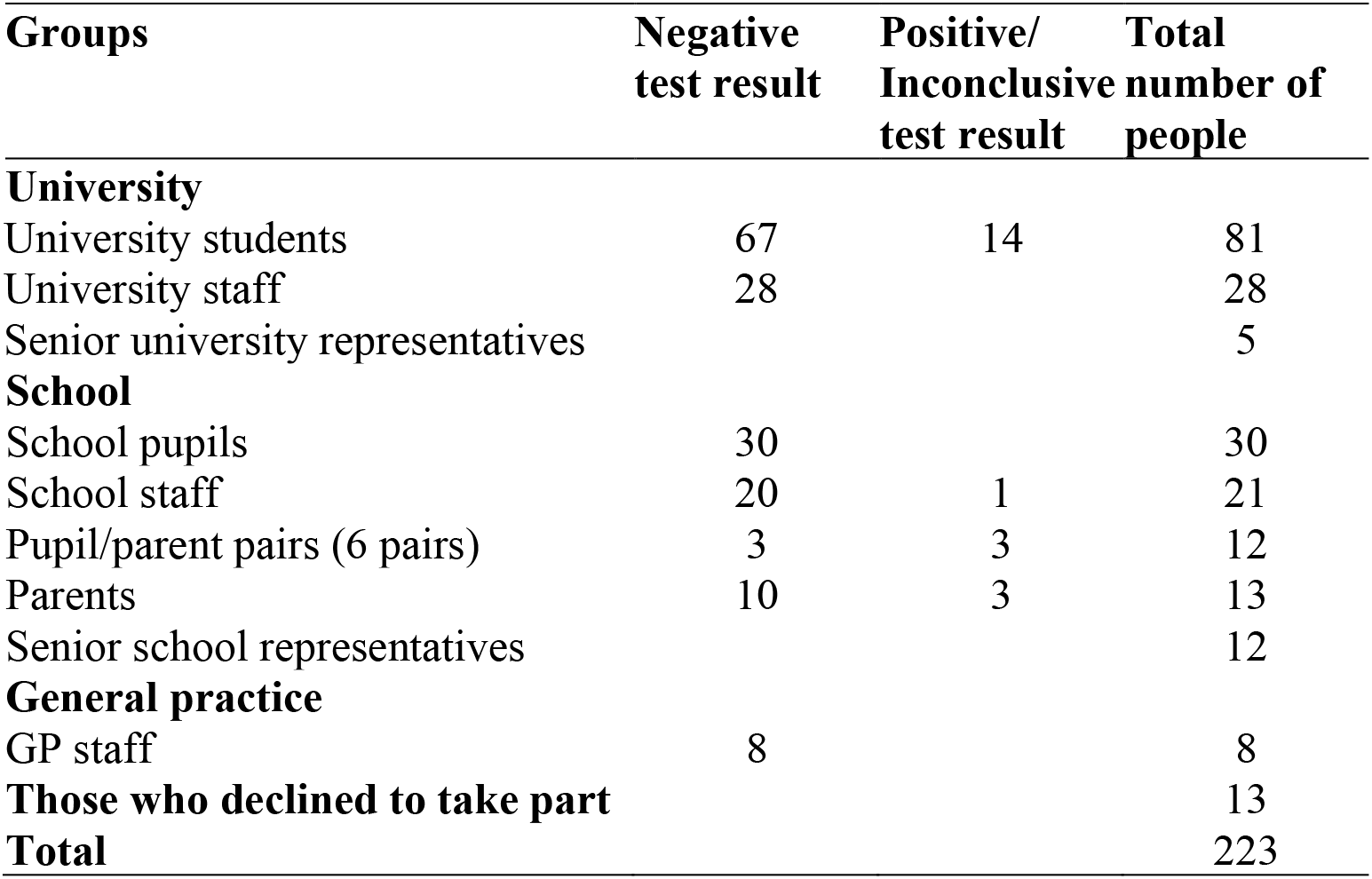
Characteristics of participants (n = 223)

| **Groups** | **Negative**<br>**test result** | **Positive/**<br>**Inconclusive**<br>**test result** | **Total**<br>**number of**<br>**people** |
| --- | --- | --- | --- |
| **University** |  |  |  |
| University students | 67 | 14 | 81 |
| University staff | 28 |  | 28 |
| Senior university representatives |  |  | 5 |
| **School** |  |  |  |
| School pupils | 30 |  | 30 |
| School staff | 20 | 1 | 21 |
| Pupil/parent pairs (6 pairs) | 3 | 3 | 12 |
| Parents | 10 | 3 | 13 |
| Senior school representatives |  |  | 12 |
| **General practice** |  |  |  |
| GP staff | 8 |  | 8 |
| **Those who declined to take part** |  |  | 13 |
| **Total** |  |  | 223 |

Focus groups were conducted using Zoom Pro and audio-recorded using Open Broadcast Software; telephone interviews were recorded using Olympus recorders and a microphone earpiece. Participants gave verbal consent for discussions to be recorded at the start of each interview and focus group. Focus groups and interviews were conducted using semi-structured discussion guides developed by experienced qualitative researchers informed by the needs of the programme team and revised iteratively in the light of insights gained. Findings from the focus groups and interviews were synthesised rapidly and added to a report, which was edited collectively by the evaluation team and continuously updated as new data and insights were generated. This report was used to provide immediate feedback to the programme team enabling them to make modifications to the running of the programme, and to offer insights to other groups in the UK developing similar testing programmes. At the end of the pilot phases of the programme, rapid analysis of the material gathered in the shared report was conducted using a framework of themes which addressed three questions:^(15)^ (i) what made people engage with the testing programme?; (ii) how could engagement with the testing programme be improved?; and (iii) what were the impacts on participants of engaging with the testing programme? Notes taken during the focus groups and interviews guided the selection of quotes which were transcribed directly from the audio-recordings.^(11, 16, 17)^ Direct quotes from participants illustrating the range of responses as they relate to the three questions are provided in Table 2. In a final phase of analysis, a public consultation activity was undertaken with public contributors hosted by the Wessex Public Involvement Network, during which our interpretation of the data was validated.

**Table 2.** Illustrative quotes.

| Theme | Quote |
| --- | --- |
| **1. What made people engage with the testing programme?** |  |
| Communication | *I would say that communication-wise from school to parents to communities - it has been strengthened... Our head teachers have been incredibly good with communication. ID46 School staff, Focus Group* |
|  | *Firstly, I didn't know what it was about. Then when I got the newsletter, it told me all about it and when it told me all about it, I was feeling more confident and more calm. ID50 School pupils, Focus Group* |
| Community | *We know that we are keeping everyone healthy....It's about our whole community, we are all in this together. ID50 School pupils, Focus Group* |
|  | *We have a WhatsApp group.. Its been good in that people have been like 'I have some glitches and stuff, I have, you know, or I'm late to deliver'.... I didn't actually see the message on my phone [test result], it was more that I heard that other people had received their message on their phone.... we have that WhatsApp group and people are quite active on that. ID03 GP staff, Interview* |
|  | *When they [the children] had all the activities, they absolutely loved that and [it] helped them to understand a lot better what's going on around them. They loved the story, the mask, the glitter and [it] made them understand what's actually going on. ID46 School staff, Focus Group* |
| Convenience | *It's quite easy. The instructions were really clear, so I understood it. ID54 Parent & Child, Interview* |
|  | *It's good that you can just walk up to campus and drop it off, and you know very quickly what the result is. ID72 University students, Focus Group* |
| **2. How could engagement with the testing programme be improved?** |  |
| Building trust | *People are a little bit wary that samples of their saliva, their nose cells, their cheek swabs are being kept. ID83 University student, Interview* |
|  | *I wouldn't want to share any data with Test and Trace at all. ID10 Decliner, Interview* |
| Extra support for testing positive | *I think my primary concern would be, obviously everyone I've come into contact with and then also income... a lot of student work is casual and obviously I can't be furloughed and sick-pay isn't the best with casual contracts. ID63 University students, Focus Group* |
|  | *They [NHS Test and Trace] were ringing, you know, nearly every single day, saying you need to stay home. Constantly ringing, constantly, literally. ID68_School parent, Interview (Child with Positive result)* |
| Increasing accessibility | *If they live a little bit further out they'd have to take public transport to get to campus which is obviously putting themselves and others at risk ... to drop off. ID80 University students, Focus Group* |
| Practical improvements | *With the labelling I did just double check it as I wasn't sure whether to stick the label over the existing label on the pot, or write it on in pen. ID44 University students, Focus Group* |
|  | *The only not so successful bit for us was my three year old really struggled to provide enough [saliva] so unfortunately we couldn't get him part of the testing programme but that was more the logistics of getting a three year old to spit. ID43 University students & staff, Focus Group* |
|  | *But it's been a case of collating the packs for staff, you know, using our time to do that. Creating emails for internal staff to say actually we are going to summarise the booklet ... it's been a bit time heavy. ID09 GP representative, Interview* |
| **3. What were the impacts on participants of engaging with the screening programme?** |  |
| Reassurance | *My partner's grandparents live quite close by. Being able to do the test and know that we didn't have it on the day that we then went to see them and could give them a hug, was an amazing thing. ID15 University staff, Interview* |
|  | *It is a huge reassurance. This empowered parents to be able to make decisions about whether to send their child to school. This head teacher has volunteered to advise other schools entering the programme to iterate how amazing this programme is in keeping communities safe, and ensuring the continuity of education. ID92 Senior school representative, Interview* |
| Pride | *I thought it [the testing programme] was a good idea and it's an honour to be part of a test trial for what could help the world. ID49 School pupil, Focus Group* |
|  | *We're really grateful for being involved and it's been a real privilege. I had a conversation with [member of staff] this morning and again they thanked me for organising it. ID90 Senior school representative, Interview* |
|  | *It's given an example to the local community of the benefit of having research-intensive, quality university on their doorstep. You can imagine 'what's the university ever done for us?'... The University has received letters particularly from schools that are a part of Phase II, just saying how much they welcome the role the University has played in making them feel safer. ID94 Senior University representative, Interview* |
| Environmental impact | *The other thing that came back from my team that they thought about was the amount of plastic that was part of the pack and no way really to recycle it. Using quite a lot of plastic as part of the process was one of the not so positive things I think. ID43 University students & staff, Focus Group* |
| Increased chance of infection | *You have got to actually touch a receptacle to put the bags in [at the drop-off point] and there was no gel or wipes or anything close by and they [friend] were a bit panicky and had to use their own stuff. ID47 University students & staff Focus Group* |
| Feelings of anxiety | *I would panic if I tested positive because I have asthma... With all the [reports] I hear, I would be intubated. I would die. That would be the end of my life. ID43 University students & staff, Focus Group* |
| Cultural beliefs | *There is language barrier at our school as well. ... Yesterday we were on the verge of finishing this lovely programme, we got the translation letters. ... We've got Urdu, Pashto, Romanian, Somali, Polish, Bengali, so we have these six translations. I think that will make a difference for next week and increase the uptake of the programme. ID46 School staff, Focus Group* |
|  | *In, especially Hong Kong right now its quite scary. If you have the saliva test or swab test right now the testing company or the government might get your DNA. Some of my friends are quite concerned about this part but because I just had to explain to them that the UK is different, they treat privacy very seriously. I try to tell them and reassure them but they are quite worried because in Hong Kong they are scared about the DNA or that the samples are being sent to China. ... But the UK might still have possibility to have accidental leaks if there are like other situations from airline companies, they have glitches in the system, some privacy just leaked out. .... Some freshers coming in this year, they are quite worried about the situation as it's been a long-standing issue there. ID48 University students & staff, Focus Group* |

### 1. What made people engage with the testing programme?

#### Communication

Participants emphasised the need for open and transparent communication from programme implementers of the reasons they should register for the programme, how to go about registering and why they should stay registered. They felt these communications should be motivating in content. To do this, they needed to address participants’ sense of community and make clear how easy and convenient the testing regime was for both those who took part and for those who were managing the programmes within their organisations. Parents, school pupils and staff described finding open communication with the programme team reassuring and therefore motivating. This open communication was led by senior representatives of the organisations taking part, who also emphasised preparation, leadership buy in and using data management systems to scale up as important to the success of the programme within their organisations. Transparent information sharing between the testing team, participants, local and national stakeholders was seen as a strength by programme implementers. Programme communications were also translated into seven languages in Phase II to increase uptake by minority ethnic groups, seen by participants as another strength.

#### Community

Participants’ decisions to take part and engage in the programme were, they felt, influenced by a pull on their sense of community. Participants from schools and GP surgeries felt this pull more strongly than participants from the University. The schools and GP surgeries were smaller, more cohesive organisations, where staff and pupils saw one another every day, spoke about the saliva testing frequently and encouraged one another to take part. This was less evident in the University. Automatic registration of all school students may also have normalised participation in the testing programme, where university students had to make the decision to register as individuals. University students suggested that taking part in the testing programme could be normalised by employing testing champions in their halls and courses to promote the programme and by using an ‘opt-out’ process for participation. School pupils and university students were offered interactive educational activities being run to increase engagement with the testing programme. Those who took part in these activities reported understanding more about the science involved in managing the pandemic, which they felt increased their motivation to protect their communities. Senior school and university representatives appreciated the role that the programme played in connecting senior community stakeholders across Southampton in a way that had not been evident before. Finding common cause through using the Saliva Testing Programme was seen as an unexpected benefit from their involvement.

#### Convenience

Most participants found the programme procedures easy and convenient; registration processes were simple, drop off points were accessible, testing instructions were clear. It perceived to be easier to carry out than nasopharyngeal PCR swab tests and test results were received quickly. They felt that making participation as convenient and easy as possible was key to increasing uptake. Parents reported that the test was simple enough for children to take responsibility for carrying out tests independently. From the perspective of the organisations, initial engagement was motivated by the cost-effectiveness of the programme and the value of having data on infection to manage outbreak hotspots by integrating programme data with data from symptomatic PCR testing, to keep the schools, the university and GP practices running.

### 2. How could engagement with the testing programme be improved?

#### Building trust

A major reason some chose not to take part in the programme was that they did not trust the Government with their data. Many of those who declined to take part in the Saliva Testing Programme were anxious about the possibility of losing control of their data when the programme passed them to NHS Test and Trace in the event of a positive test. The local NHS Foundation Trust and its partnership with the University and Southampton City Council, however, was trusted; it was seen to have scientific integrity, and as a local organisation, was felt to be answerable to the Southampton community in a way NHS Test and Trace was not. Some suggested that they would have been more likely to take part if the programme was run solely by local organisations. Participants and decliners were clear that building trust was necessary to improve engagement in the programme. They suggested that this would be helped by receiving more directed information from credible sources such as the university and Southampton City Council about the rationale for and design of the programme, about data protection and the accuracy of the tests, and about the progress of the programme. This information would increase transparency and help dispel myths, particularly about the accuracy of the saliva test. One reason some people declined to take part in the programme was a concern about the personal consequences of a false-positive result.

#### Extra support for testing positive

Those who had experienced a positive test result asked for more efficient data management by the testing programme, NHS Test and Trace and their general practice, and more coordinated messaging. Participants requested more personalised support for those testing positive and having therefore to self-isolate. This included financial aid if unable to work, receiving food and medication supplies and mental health support. Some felt, however, that there were too many support calls from NHS Test and Trace for those testing positive. Participants were particularly worried about the possibility of spreading the virus to others whilst they waited for a test result, and feared the stigma of testing positive, which suggests that they would need reassurance and social support in dealing with a positive test result. Fear of a positive test result was enough to make some decline to take part; they were concerned that if they had to isolate they would lose income, their employer would be unsympathetic and that a history of infection with the virus might affect their ability to get a mortgage and life-insurance. These people preferred not to know their viral status.

#### Increasing accessibility

Both decliners and participants recommended making the programme more inclusive and accessible to a wider group of those eligible to take part. Drop-off points for saliva samples that catered for people living outside Southampton and more frequent, convenient and better sign-posted drop-off points within Southampton would have reduced the distances people had to travel. Some suggested postal deliveries and returns. Comprehensive translation of all programme communications would have increased uptake and engagement of those from minority ethnic communities.

#### Practical improvements

Many improvements suggested by participants were incorporated into the programme as it evolved (see Box 1). A small minority of participants, especially parents of younger children, had issues producing enough saliva, which may have produced an inconclusive result. They suggested the tubes be marked with a clear indicator of the amount necessary. Some decliners missed the University registration email and recommended putting key information highlighted at the top of emails. Senior representatives of organisations involved were clear that the programme gave their staff added responsibilities and added to their workload. Whilst accepting that this extra work was in a good cause, some suggested a ‘toolkit’ of instructions and tips for those implementing the programme to help manage the expectations of both staff and participants.

### 3. What were the broader consequences for participants of engaging with the testing programme?

Participants reflected on the positive and negative impacts of participating in the testing programme. Positive outcomes included:

#### Reassurance

They generally felt reassured by knowing their viral status and expressed a sense of relief and reduced feelings of anxiety when they tested negative. They appreciated knowing they were not spreading the virus, felt that this enabled a “near normal” life to continue and were more confident going to school or work and visiting vulnerable family and friends. This was perceived to be an incentive for taking part in the programme.

#### Pride

Participants expressed pride in knowing that they were contributing to a programme that was part of the national effort to manage the pandemic. Some viewed this as a privilege and others were excited. Southampton university students spoke of being envied by those from other universities and felt that the testing programme made the university attractive to prospective students. Senior university representatives spoke of the reputational benefit to the organisation of having provided a testing programme for the local community.

Participants expressed a number of concerns

#### Environmental impact

Some were concerned about the amount of plastic in testing kits and the environmental impact of an expansion of the testing programme.

#### Increased chance of infection

Concern was expressed about the potential of those who received a negative test result to become less vigilant in applying social-distancing and hygiene measures.

They were also worried that the test kit drop-off points were sites of potential infection; prior Health and Safety risk assessments had however ensured that these carried no or minimal risk.

#### Feelings of anxiety

Participants reported feelings of anxiety whilst waiting for their test results, worrying about the personal consequences of having to self-isolate or of having unknowingly passed the virus on to others whilst awaiting their test result. Others were anxious about the possibility of the testing programme ending.

#### Cultural barriers

Some participants were concerned that aspects of culture were a barrier to participation. These included language barriers and a cultural mistrust in central government. Those who had lived in countries where governments were believed to misuse personal data were more anxious about taking part in the Saliva Testing Programme because of the perceived threat to their privacy. Those who needed the testing programme most may in this way have been prevented from taking part.

## Conclusions

Individual participants and organisational representatives provided valuable insights into the experience of engaging with the Southampton COVID-19 Saliva Testing Programme, leading to changes in programme processes and communications, which undoubtedly increased engagement and adherence to programme requirements. They also made important observations about the benefits they experienced from engaging with the testing programme, some of which they suggested should be emphasised to potential participants by those currently establishing testing programmes. One of these benefits was increased knowledge about the science behind management of the pandemic that school and university students gained from the educational engagement activities. As well as providing insights, it was clear from the focus groups and interviews that the process of seeking feedback from participants also increased the engagement of local people with the testing programme.

Involving local people and organisations in the development and piloting of programmes in this way may well have been important in ensuring community buy-in.^(18)^ They appreciated the chance to contribute to this programme, which they saw to be of national importance. The process of holding the conversations reported in this paper may also have helped to address two key underlying issues affecting engagement with the testing programme: the need for trust between participants and testing programmes, and the role of ‘collective efficacy’ within organisations. Local organisations, such as schools and the university were seen to be answerable to local people and hence more trustworthy than more national ‘faceless’ and private organisations such as NHS Test and Trace. Collective efficacy is a group’s shared belief in its capability to organise and execute actions required to achieve goals,^(19)^ and a sense of collective efficacy appeared to be stronger in the smaller, more cohesive organisations such as the schools and GP practices than in the University, for example. Schools and GP practices had staff and student communities within which members through daily conversation and mutual encouragement made regular testing the ‘social norm’. The educational engagement activities appeared to play an important role in this too. Students’ increased knowledge appeared to make them more engaged with the programme and motivated by the need to protect their communities, as well as making them feel like they had more agency in controlling the spread of the virus and its damaging consequences. Schools and the university were thus building collective efficacy for regular testing.

A rapid qualitative evaluation enabled real-time feedback of insights into the testing programme, improving the participants’ experience and was therefore likely to increase uptake of testing. Insights gained were also used to inform university, schools, and city-wide strategies for managing the pandemic and to feed-back to national and local government.^(20)^ Figure 1 illustrates the way in which the rapid qualitative evaluation influenced Saliva Testing Programme development and wider testing strategy and Box 1 details the modifications made to the programme based on feedback gathered through the interviews and focus group discussions. This study has also produced actionable recommendations for improving engagement with COVID-19 testing programmes across the UK. See Box 2 for details of these recommendations.

**Figure 1.**
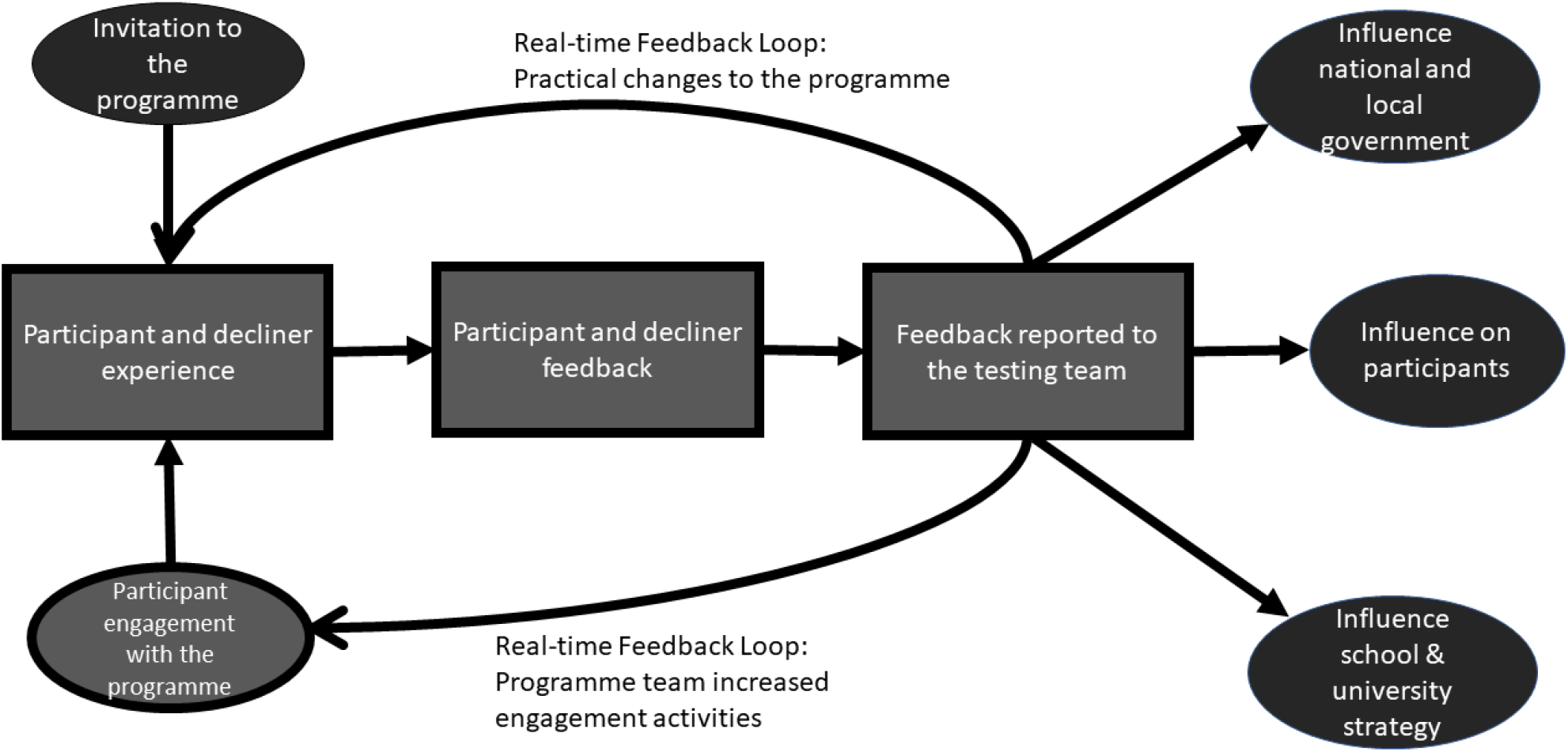
How participants’ experiences informed the Southampton COVID-19 Saliva Testing Programme wider strategy.

### Strengths and limitations

The speed with which insights were needed to optimise the testing programme meant that a full and rigorous traditional qualitative analysis of the data was not possible. Steps taken to ensure that our findings were robust included regular team meetings to discuss interpretations, checking these interpretations with participants in focus groups and interviews and consultation with experts from a wide range of backgrounds. These steps are part of an established process for conducting rapid qualitative evaluation.^(11-17)^ Accessing those who declined to take part in the programme was challenging, but those who were consulted offered important insights that informed significant programme development. Future evaluations of testing programmes should explore participants’ experiences of indirect NHS Test and Trace contact tracing and household isolation as these are key components of the effectiveness of population testing.

#### Box 1

**Figure 1 Key and Modifications made to the Saliva Testing Programme as a result of participant feedback:**

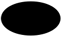 Programme inputs and outputs

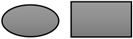 Participant experiences of the programme

**Modifications to the Saliva Testing Programme:**

1. Resolved initial technical hurdles in the registration process and moved towards a more robust yet simple registration process in Phase II to reduce the barriers of registering;
2. Created clear and simple instructions, which were translated into seven languages, to reduce the number of inconclusive saliva test results;
3. Designed smaller labels to stick to the test pots to reduce the number of people placing them incorrectly and increasing the risk of receiving an inconclusive result;
4. Testing team communicated the test’s accuracy and the progress of the programme in a weekly newsletter to schools and through emails to university staff and students;
5. Educational engagement activities were offered to university students to increase their involvement in the programme;
6. Commitment has been made to reduce the amount of plastic and to recycle containers in the next phase of the programme;
7. Post-boxes rather than team members placed at sample drop-off points to reduce possibility of transmission;
8. Increased number of drop-off points in Southampton to reduce the travel time for participants submitting their tests;
9. ‘Toolkit’ created for programme implementers to support preparation and roll-out for the next phase of the programme.

#### Box 2

**Recommendations from the Southampton Saliva Testing Programme to ensure mass engagement in testing for COVID-19**

1. Testing should be delivered through local organisations to both increase trust in the testing programme but also to promote collective efficacy;
2. Communications about testing should be clear, consistent and appeal to individual’s sense of community and altruism to motivate people to take part in the programme;
3. Creative and fun educational activities should be used to improve knowledge and understanding of the virus, so increasing motivation to protect each other and sense of agency in managing consequences of the pandemic;
4. Participants and local organisers should be involved in designing their programme, and should be engaged in providing continuous feedback on the testing experience to enable real-time programme modifications;
5. Local organisations involved in delivering testing should be enabled to connect with one another to share best practice and create a local testing culture;
6. Those testing positive should be supported financially, psychologically, with food and medication and provided with reassurance and advice about how to minimise the possibility of transmission of infection to others;
7. Testing should be made as convenient for participants as possible and all communications need to be in multiple languages as well as appropriate for children and young people;
8. Thought needs to be given to making testing kits and processes as environmentally sustainable as possible.

## Data Availability

Data is available on request.

## Acknowledgements

The authors would like to thank the participants for their time and interest in the interviews and focus groups. We would also like to thank the Southampton COVID-19 Testing Pilot Programme team for their help with recruitment and data collection, the University of Southampton, Southampton City Council and University Hospital Southampton for their support with evaluating the programme, and the Department of Health and Social Care for funding the project.

